# From population to neuron: exploring common mediators for metabolic problems and mental illnesses

**DOI:** 10.1101/2020.09.13.20183525

**Authors:** Yoichiro Takayanagi, Koko Ishizuka, Thomas M. Laursen, Hiroshi Yukitake, Kun Yang, Nicola G. Cascella, Shuhei Ueda, Akiko Sumitomo, Zui Narita, Yasue Horiuchi, Minae Niwa, Akiko Taguchi, Morris F. White, William W. Eaton, Preben B. Mortensen, Takeshi Sakurai, Akira Sawa

**Affiliations:** Department of Mental Health, Johns Hopkins University Bloomberg School of Public Health, Baltimore, MD, USA; Department of Psychiatry, Johns Hopkins University School of Medicine, Baltimore, MD, USA; Department of Neuroscience, Johns Hopkins University School of Medicine, Baltimore, MD, USA; Department of Biomedical Engineering, Johns Hopkins University School of Medicine, Baltimore, MD, USA; Department of Genetic Medicine, Johns Hopkins University School of Medicine, Baltimore, MD, USA; National Centre for Register-Based Research, Department of Economics and Business Economics, Aarhus University, Denmark; The Lundbeck Foundation’s Initiative for Integrative Research, iPSYCH; Center for Integrated Register-based Research at Aarhus University, CIRRAU; Medical Innovation Center, Kyoto University Graduate School of Medicine, Japan; Department of Integrative Aging Neuroscience, National Center for Geriatrics and Gerontology, Japan; Division of Endocrinology, Department of Medicine, Boston Children’s Hospital, Harvard Medical School, Boston, MA, USA

**Author notes:** Department of Neuropsychiatry, University of Toyama Graduate School of Medicine and Pharmaceutical Sciences, Japan (Yoichiro Takayanagi). Equal contribution (Yoichiro Takayanagi, Koko Ishizuka). Corresponding authors: Contact author: Akira Sawa, M.D. 600 N Wolfe Street, Meyer 3-166, Baltimore, MD, USA.

## Abstract

Major mental illnesses such as schizophrenia (SZ) and bipolar disorder (BP) frequently accompany metabolic conditions, but their relationship is still unclear, in particular at the mechanistic level. We implemented an approach of “from population to neuron”, combining population-based epidemiological analysis with neurobiological experiments using cell and animal models based on a hypothesis built from the epidemiological study. We characterized high-quality population data, olfactory neuronal cells biopsied from patients with SZ or BP, and healthy subjects, as well as mice genetically modified for insulin signaling. We accessed the Danish Registry and observed (1) a higher incidence of diabetes in people with SZ or BP and (2) higher incidence of major mental illnesses in people with diabetes in the same large cohort. These epidemiological data suggest the existence of common pathophysiological mediators in both diabetes and major mental illnesses. We hypothesized that molecules associated with insulin resistance might be such common mediators, and then validated the hypothesis by using two independent sets of olfactory neuronal cells biopsied from patients and healthy controls. In the first set, we confirmed an enrichment of insulin signaling-associated molecules among the genes that were significantly different between SZ patients and controls in unbiased expression profiling data. In the second set, olfactory neuronal cells from SZ and BP patients who were not pre-diabetic or diabetic showed reduced IRS2 tyrosine phosphorylation upon insulin stimulation, indicative of insulin resistance. These cells also displayed an upregulation of IRS1 protein phosphorylation at serine-312 at baseline (without insulin stimulation), further supporting the concept of insulin resistance in olfactory neuronal cells from SZ patients. Finally, *Irs2* knockout mice showed an aberrant response to amphetamine, which is also observed in some patients with major mental illnesses. The bi-directional relationships between major mental illnesses and diabetes suggest that there may be common pathophysiological mediators associated with insulin resistance underlying these mental and physical conditions.

## INTRODUCTION

Epidemiological studies have provided important working hypotheses on disease pathology in many areas of medicine, including psychiatry [1]. For example, a higher incidence of schizophrenia (SZ) in individuals born during winter has hinted that immune-inflammatory disturbances during early development induced by exposure to infectious agents *in utero* may be involved in SZ pathogenesis [2, 3].

SZ and bipolar disorder (BP) are major mental illnesses affecting about 2% of the population in many countries. The dichotomy of these two disorders, delineated by Kraepelin, has been a principle of modern psychiatry [4], but recent genetic data support that these two disorders share common genetic risk factors [5]. Several population-based longitudinal studies have shown that patients diagnosed with SZ or BP have a higher risk of developing diabetes compared with the general population [6-10]. However, given that antipsychotic and antidepressant medications can elicit metabolic problems [11], it is unclear whether these epidemiological observations stem primarily from intrinsic vulnerability in patients with SZ or BP to develop metabolic conditions, or if they are a reflection of medication-induced secondary effects.

Glucose intolerance is observed in many individuals with major mental illnesses, even in drug-naïve psychotic patients [12-14]. These results imply that the higher incidence of diabetes in SZ and BP patients may be because of an intrinsic vulnerability to metabolic disturbance in these patients. Such intrinsic changes may predispose patients with major mental illnesses (SZ and BP) to diabetes, and (vice versa) those diagnosed with diabetes to major mental illnesses. Thus, we hypothesized that the incidence of developing major mental illnesses (e.g., SZ and BP) would be higher in patients already diagnosed with diabetes than in non-diabetic subjects. If this is the case, both major mental illnesses and diabetes would increase risk for the other in a bi-directional manner. As far as we are aware, no study has addressed such a bi-directional relationship in the same cohort.

In the present study, we conducted a population-based prospective epidemiological analysis using the Danish Psychiatric Registry and Danish National Diabetes Registry to address the relationship between diabetes and major mental illnesses (SZ and BP). We indeed identified a bi-directional relationship between diabetes and major mental illnesses in the same cohort. Based on this information, we hypothesized the existence of common pathophysiological mediators in both diabetes and major mental illnesses, and further hypothesized that molecules associated with insulin resistance might be common mediators. We then validated the hypothesis by using two independent sets of olfactory neuronal cells biopsied from patients and healthy controls, and obtained evidence in accordance with this notion at both gene expression and protein levels. Furthermore, in mice deficient in *Irs2*, we observed dopamine-associated behavioral changes relevant to these mental conditions.

## MATERIALS AND METHODS

### The Danish Registry and its analysis

The Danish Civil Registration System covers all residents in Denmark and can identify individuals among all national registers, enabling precise linkage [15]. We used the Danish Psychiatric Registry containing all psychiatric hospitalizations since 1977 and all psychiatric contacts at outpatient services since 1995 [16], as well as the Danish National Diabetes Registry monitoring the occurrence of diabetes since 1995 [17]. All individuals who were alive in Denmark on January 1, 1997 were included in our study. This study was approved by the Danish Data Protection Agency, Statistics Denmark, and the Danish National Board of Health in which informed consent from each subject is not legally required for registry-based studies.

Psychiatric diagnosis was performed by psychiatrists who treated those individuals. The codes used to identify cohort members with SZ or BP and the criteria used to identify members with diabetes are summarized in **Supplemental Tables 1** and **2**, respectively. The incidence of the disease of interest (i.e., diabetes, SZ, or BP) in each population is summarized in **Supplemental Table 3**.

We used a log-linear Poisson regression with the GENMOD procedure using the SAS program (version 9.2, SAS Institute, Inc.). This method estimates a Cox regression, and calculates the incidence rate that is described as relative risk (RR). RRs were adjusted for the following potential confounders: calendar year, age group, gender, parental family history of SZ, parental family history of BP, parental family history of diabetes, history of alcohol abuse, and degree of urbanicity at birth. Degree of urbanization was defined as Capital, Capital Suburb, Provincial City, Provincial Town, and Rural Area. Age, calendar year, psychiatric diagnosis, and diagnosis of diabetes were treated as time-dependent variables, while all others were treated as fixed variables. Age was categorized into 10-year intervals. Calendar year was categorized into 1-year intervals for 1997 and into 2-year intervals thereafter. We included only persons aged 15 years or older when analyzing the risk of major mental illnesses in the diabetic population, since the incidence of SZ and BP is very small in children younger than that age. We also linked the data to the Danish National Registry of Patients for additional analyses to examine potential confounding by differences in the subtypes of diabetes (i.e., types 1 and 2). We used the subdivision of diabetes only for supplemental analyses, because although the diagnosis in the Danish National Registry of Patients is based on the Danish version of the World Health Organization International Classification of Diseases, tenth edition (ICD 10), other indicators for the diagnosis of type 1 diabetes (e.g., immunological tests) were not considered.

### Analysis of gene expression in human olfactory neuronal cells (the PsychENCODE dataset)

We conducted literature mining of the genes that were differentially expressed between SZ patients and healthy controls (FDR < 0.05) in olfactory neuronal cells in the PsychENCODE dataset [18]. Genes were annotated either insulin-related or -unrelated based on the criteria that more than 100 publications mentioned its gene symbol and insulin at the same time in the title or abstract in PubMed publications. NCBI Entrez Programming Utilities (E-utilities) was used to perform a batch search for all human genes. Chi-square test for the following 2×2 contingency table was performed to evaluate whether or not insulin-related genes were enriched in these differentially expressed genes.

### Analysis of IRS1/2 phosphorylation in human olfactory neuronal cells (the Johns Hopkins cohort)

In the Johns Hopkins Schizophrenia Center, olfactory neuronal cells were prepared from nasal biopsied tissues from patients and healthy controls, according to published protocols [19, 20]. In the present study, we used neuronal cells from patients with SZ (N=15), BP (N=11), and healthy control subjects (N=15). Due to vulnerability of these primary cells to freeze-thaw process, cells from 1 heathy control and 2 SZ were not available in the experiments for IRS1. These patients were drawn from the clinical population at the Johns Hopkins Medical Institutions. Healthy controls were recruited through advertisements placed in local papers or leaflets left on hospital boards. All subjects gave informed consent that had been approved by the Johns Hopkins Hospital IRB.

To examine the cellular response to insulin, we used a published protocol with some modifications [21]. In brief, following serum starvation for 48 h in D-MEM/F-12 medium (Invitrogen), the cells were stimulated for 10 min with 10 nM of insulin (Sigma) before being harvested. The level of phospho-IRS2 was assessed by immuno-precipitation followed by Western blotting, according to an established protocol with some modifications [21-23]. In brief, cells were lysed with RIPA buffer (Cell Signaling Technologies) together with protease inhibitors (cOmplete; Roche) and phosphatase inhibitors (Thermo Scientific). Cell supernatant obtained by centrifugation at 15,000 rpm for 20 min at 4°C was used for immunoprecipitation with an anti-IRS2 antibody (Cell Signaling Technologies). Immunoprecipitates were analyzed by SDS-PAGE followed by Western blotting with an anti-pan phospho-tyrosine antibody, 4G10 (Millipore). The level of phospho-IRS1 was assessed by Western blotting with an anti-phospho-IRS1 (at human serine-312) antibody (Cell Signaling Technologies). Cell supernatant was obtained by the same protocol as used for the IRS2 experiments.

Signal detection was carried out with LAS 1000, and the signal intensity was quantified with Image J software. Statistical analysis was performed using one-way analysis of variance (ANOVA) with Bonferroni’s multiple comparison test or Kruskal-Wallis analysis followed by Dunn’s test.

### Behavioral studies of *Irs2* knockout mice

*Irs2* knockout (KO) mice have been described previously [24]. Mice were maintained at the Medical Innovation Center of Kyoto University Graduate School of Medicine with the C57Bl6/J genetic background by mating heterozygotes. All mice were group housed after weaning and throughout the behavioral testing, and kept under a 12 h light /12 h dark cycle with food and water provided *ad lib*. We used male homozygous (KO) mice and their wild type (WT) littermates for analysis. We followed the sample size and behavioral assay protocols in our past publications [20, 25-28].

We used two cohorts: For the first cohort, the following behavioral tests were performed day by day in 7 weeks of age: weight check and open field; Y-maze; three-chamber social interaction; elevated plus maze; forced swim; and weight check and rotarod. The second cohort was used for methamphetamine challenge (postnatal days 48-52): we measured their body weight and performed open field for the first 60 min following an intraperitoneal saline injection, and then for another 120 min following an intraperitoneal methamphetamine injection (1 mg/kg). Using total distance traveled during the last 10 min before methamphetamine injection as baseline, we assessed the fold change relative to the baseline activity after methamphetamine injection [29]. Behaviors were recorded and analyzed automatically to keep objectiveness.

All data are presented as the mean with standard error of the mean. Statistical analysis was performed using univariate or repeated measures ANOVA, or F-test with either Student’s t or Welch’s t test. For measurements of methamphetamine-induced hyperactivity, statistical significance was determined using two-way ANOVA and subsequent Sidak’s post hoc analysis for multiple comparisons.

## RESULTS

### Higher risk of developing diabetes after the onset of major mental illnesses (SZ and BP)

First, we addressed whether patients diagnosed with SZ or BP have a higher risk of later developing diabetes compared with the general population. We took advantage of the Danish Civil Registration System that enabled us to follow individuals with diseases of interest in the entire Danish population. We linked the registers in the Danish Psychiatric Registry with those in the Danish National Diabetes Registry, both described in detail in the section on Materials and Methods. We performed a prospective study from January 1, 1997 to one of the following time points: onset of the disease of interest, relocation out of Denmark, or December 31, 2009, whichever came earliest.

We compared the incidence of diabetes among four groups, namely subjects with SZ, without SZ, with BP, and without BP, as rate per 1000 person-years (**Supplemental Table 3a**). Then, we calculated the incidence-rate ratios between groups with and without disorders, which could be interpreted as the RR for the disorders. Analyses showed that the incidence of diabetes was higher in both patients with SZ [adjusted RR = 2.16, 95% confidence interval (CI) = 2.03-2.29 for female, adjusted RR = 1.78, 95% CI = 1.68-1.88 for male] and patients with BP (adjusted RR = 1.63, 95% CI = 1.53-1.74 for female, adjusted RR = 1.36, 95% CI = 1.26-1.48 for male) compared with the general population (**Fig. 1a**). The full models of the log linear regression analyses are summarized in **Tables 1a** and **1b**. These are consistent with several previous reports [6-9]. Although women were less likely to develop diabetes than men in the population registered in the depository (adjusted RR = 0.83, 95% CI = 0.82-0.84), women were at higher risk of developing diabetes when they also had SZ or BP.

**Fig. 1.**
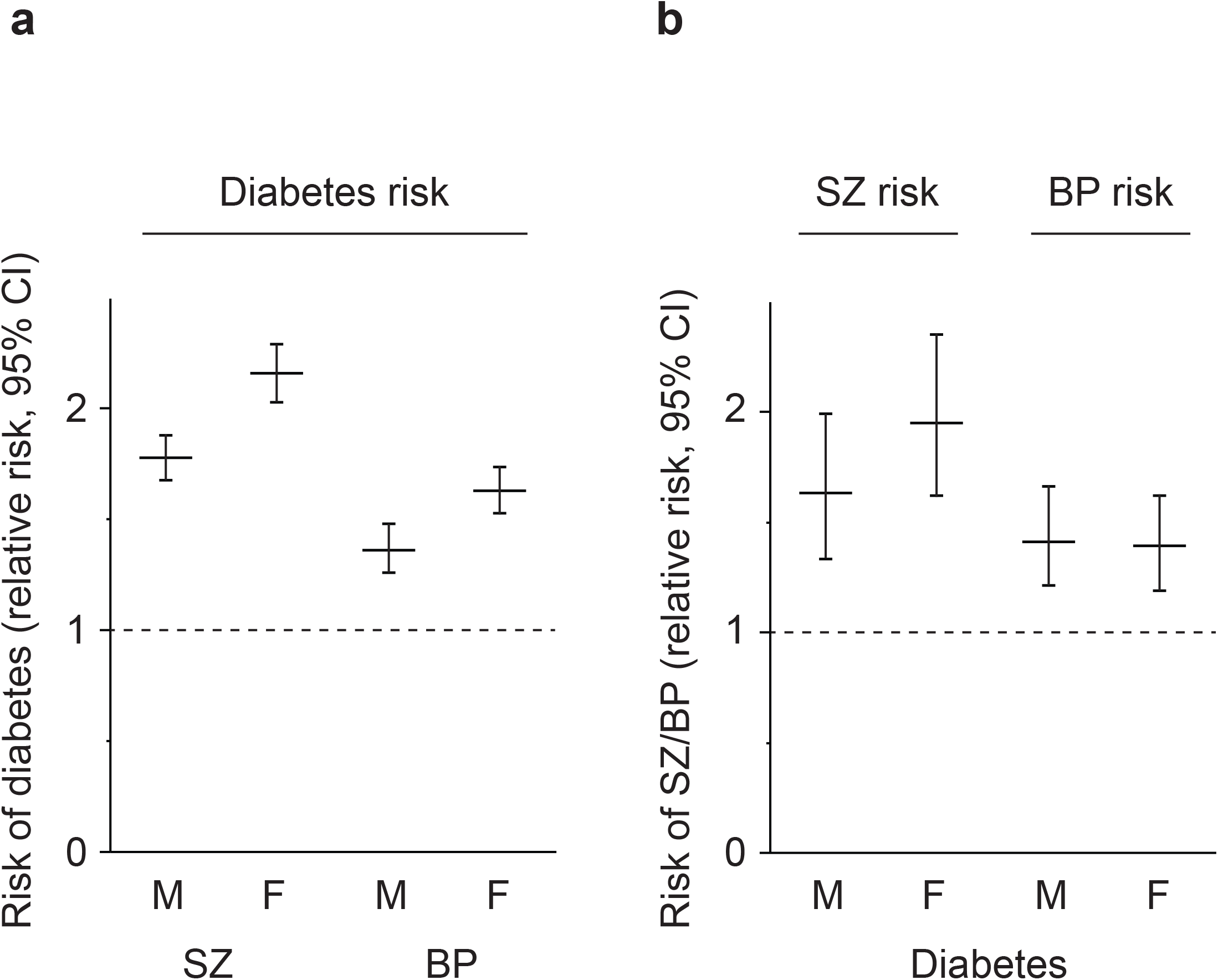
Relations between risk of diabetes and severe mental illnesses. (**a**) The risk of diabetes in patients with schizophrenia (SZ) and bipolar disorder (BP) is estimated by analysis of the National Registry in Denmark. Lines indicate relative risk, and bars indicate ranges with 95% confidence interval (CI). Data are shown separately for each gender. Females showed higher incidence of SZ and BP than males in the National Registry. (**b**) The risk of SZ and BP in patients with diabetes is estimated by analysis of the National Registry in Denmark. Lines indicate relative risk, and bars indicate ranges with 95% CI. Data are shown separately for each gender. Note that there is no gender difference in the incidence of diabetes in the National Registry.

**Table 1a.**
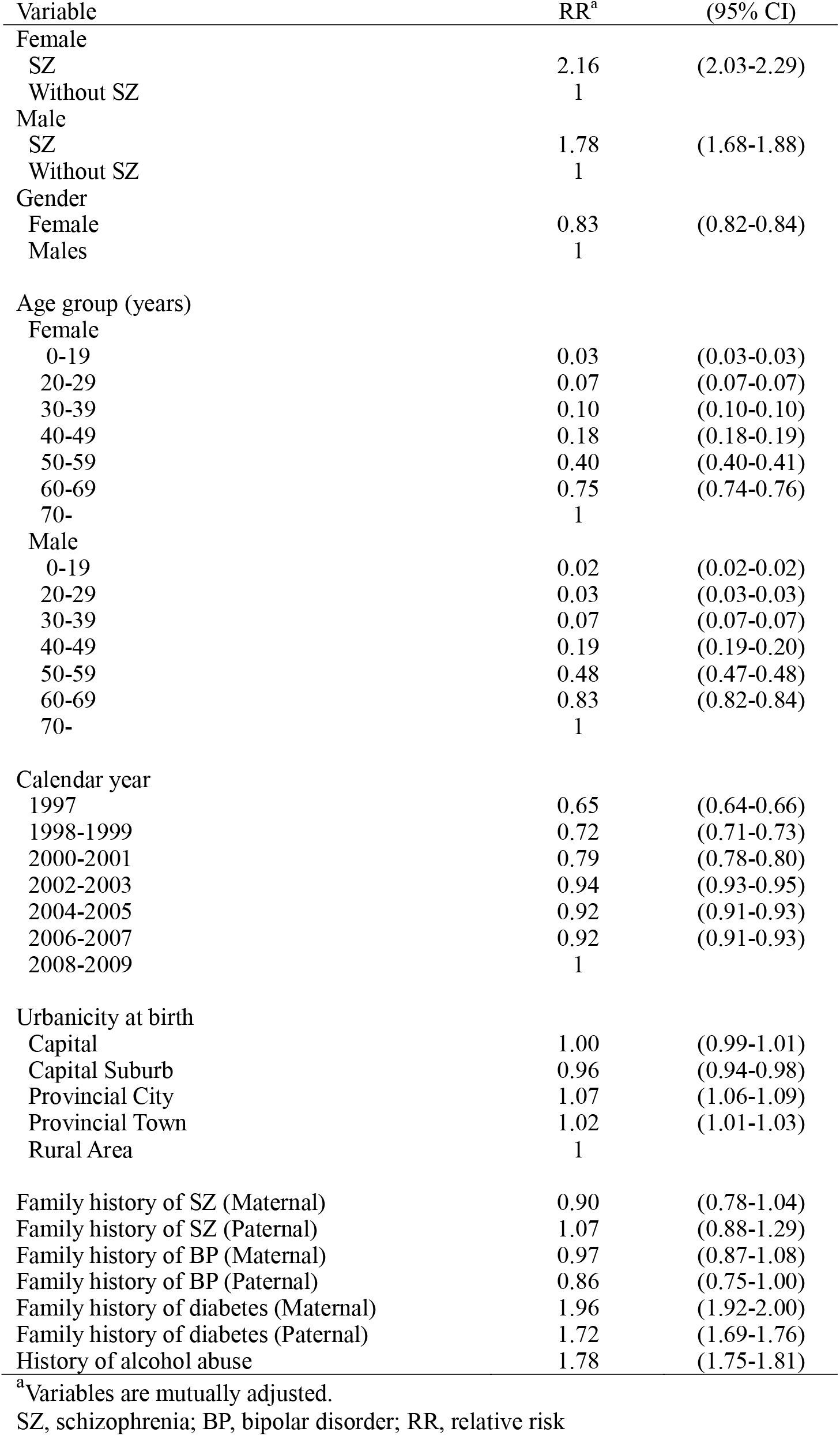
Full models of log linear regression analysis for risk of diabetes in patients with schizophrenia (SZ)

| Variable | RR<sup>a</sup> | (95% CI) |
| --- | --- | --- |
| Female |  |  |
| SZ | 2.16 | (2.03-2.29) |
| Without SZ | 1 |  |
| Male |  |  |
| SZ | 1.78 | (1.68-1.88) |
| Without SZ | 1 |  |
| Gender |  |  |
| Female | 0.83 | (0.82-0.84) |
| Males | 1 |  |
| Age group (years) |  |  |
| Female |  |  |
| 0-19 | 0.03 | (0.03-0.03) |
| 20-29 | 0.07 | (0.07-0.07) |
| 30-39 | 0.10 | (0.10-0.10) |
| 40-49 | 0.18 | (0.18-0.19) |
| 50-59 | 0.40 | (0.40-0.41) |
| 60-69 | 0.75 | (0.74-0.76) |
| 70- | 1 |  |
| Male |  |  |
| 0-19 | 0.02 | (0.02-0.02) |
| 20-29 | 0.03 | (0.03-0.03) |
| 30-39 | 0.07 | (0.07-0.07) |
| 40-49 | 0.19 | (0.19-0.20) |
| 50-59 | 0.48 | (0.47-0.48) |
| 60-69 | 0.83 | (0.82-0.84) |
| 70- | 1 |  |
| Calendar year |  |  |
| 1997 | 0.65 | (0.64-0.66) |
| 1998-1999 | 0.72 | (0.71-0.73) |
| 2000-2001 | 0.79 | (0.78-0.80) |
| 2002-2003 | 0.94 | (0.93-0.95) |
| 2004-2005 | 0.92 | (0.91-0.93) |
| 2006-2007 | 0.92 | (0.91-0.93) |
| 2008-2009 | 1 |  |
| Urbanicity at birth |  |  |
| Capital | 1.00 | (0.99-1.01) |
| Capital Suburb | 0.96 | (0.94-0.98) |
| Provincial City | 1.07 | (1.06-1.09) |
| Provincial Town | 1.02 | (1.01-1.03) |
| Rural Area | 1 |  |
| Family history of SZ (Maternal) | 0.90 | (0.78-1.04) |
| Family history of SZ (Paternal) | 1.07 | (0.88-1.29) |
| Family history of BP (Maternal) | 0.97 | (0.87-1.08) |
| Family history of BP (Paternal) | 0.86 | (0.75-1.00) |
| Family history of diabetes (Maternal) | 1.96 | (1.92-2.00) |
| Family history of diabetes (Paternal) | 1.72 | (1.69-1.76) |
| History of alcohol abuse | 1.78 | (1.75-1.81) |

<sup>a</sup>Variables are mutually adjusted.
SZ, schizophrenia; BP, bipolar disorder; RR, relative risk

**Table 1b.** Full models of log linear regression analysis for risk of diabetes in patients with bipolar disorder (BP)

| Variable | RR<sup>a</sup> | (95% CI) |
| --- | --- | --- |
| Female |  |  |
| BP | 1.63 | (1.53-1.74) |
| Without BP | 1 |  |
| Male |  |  |
| BP | 1.36 | (1.26-1.48) |
| Without BP | 1 |  |
| Gender |  |  |
| Female | 0.83 | (0.82-0.84) |
| Males | 1 |  |
| Age group (years) |  |  |
| Female |  |  |
| 0-19 | 0.03 | (0.03-0.03) |
| 20-29 | 0.07 | (0.07-0.07) |
| 30-39 | 0.10 | (0.10-0.10) |
| 40-49 | 0.18 | (0.18-0.19) |
| 50-59 | 0.40 | (0.40-0.41) |
| 60-69 | 0.75 | (0.74-0.76) |
| 70- | 1 |  |
| Male |  |  |
| 0-19 | 0.02 | (0.02-0.02) |
| 20-29 | 0.03 | (0.03-0.03) |
| 30-39 | 0.07 | (0.07-0.07) |
| 40-49 | 0.19 | (0.19-0.20) |
| 50-59 | 0.48 | (0.47-0.49) |
| 60-69 | 0.83 | (0.82-0.84) |
| 70- | 1 |  |
| Calendar year |  |  |
| 1997 | 0.65 | (0.64-0.66) |
| 1998-1999 | 0.72 | (0.71-0.73) |
| 2000-2001 | 0.79 | (0.78-0.80) |
| 2002-2003 | 0.94 | (0.93-0.95) |
| 2004-2005 | 0.92 | (0.91-0.93) |
| 2006-2007 | 0.92 | (0.91-0.93) |
| 2008-2009 | 1 |  |
| Urbanicity at birth |  |  |
| Capital | 1.00 | (0.99-1.01) |
| Capital Suburb | 0.96 | (0.94-0.98) |
| Provincial City | 1.07 | (1.06-1.09) |
| Provincial Town | 1.02 | (1.01-1.03) |
| Rural Area | 1 |  |
| Family history of SZ (Maternal) | 0.93 | (0.80-1.07) |
| Family history of SZ (Paternal) | 1.09 | (0.90-1.32) |
| Family history of BP (Maternal) | 0.97 | (0.87-1.08) |
| Family history of BP (Paternal) | 0.86 | (0.74-1.00) |
| Family history of diabetes (Maternal) | 1.97 | (1.93-2.01) |
| Family history of diabetes (Paternal) | 1.72 | (1.69-1.76) |
| History of alcohol abuse | 1.80 | (1.77-1.83) |

<sup>a</sup>Variables are mutually adjusted.
SZ, schizophrenia; BP, bipolar disorder; RR, relative risk

### Higher risk of developing major mental illnesses (SZ and BP) after the onset of diabetes

We next addressed the risk of developing (or diagnosed with) SZ or BP between patients with diabetes and the general population (**Supplemental Tables 3b** and **3c**). We employed the same strategy that was used to validate higher risk of developing diabetes after the onset of SZ or BP.

We observed an increased risk of developing SZ or BP in the diabetic population compared to the general population (for SZ, adjusted RR = 1.95, 95% CI = 1.62-2.35 for female, adjusted RR = 1.63, 95% CI = 1.33-1.99 for male; for BP, adjusted RR = 1.39, 95 % CI = 1.19-1.62 for female, adjusted RR = 1.41, 95% CI = 1.21-1.66 for male) (**Fig. 1b**). The full models of the regression analyses are summarized in **Tables 2a** and **2b**. In the registered population, women had a higher risk of developing both SZ (adjusted RR = 2.61, 95% CI = 1.74-3.89) and BP (adjusted RR = 1.50, 95% CI = 1.26-1.79) than men. Women with diabetes also had a higher risk of developing SZ, but not BP, than men.

**Table 2a.**
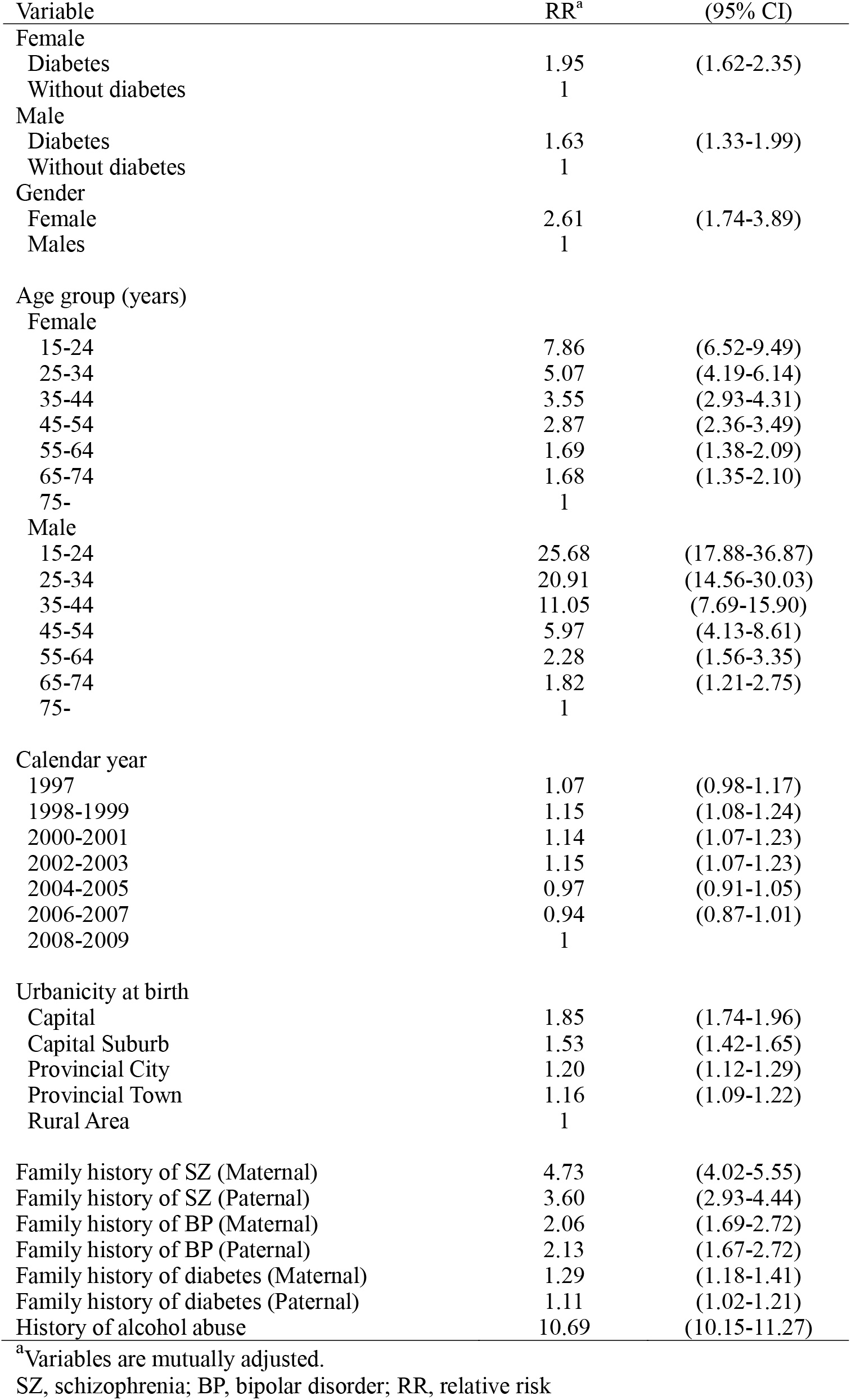
Full models of log linear regression analysis for risk of SZ in the diabetic population.

**Table 2b.** Full models of log linear regression analysis for risk of BP in the diabetic population.

| Variable | RR<sup>a</sup> | (95% CI) |
| --- | --- | --- |
| Female |  |  |
| Diabetes | 1.39 | (1.19-1.62) |
| Without diabetes | 1 |  |
| Male |  |  |
| Diabetes | 1.41 | (1.21-1.66) |
| Without diabetes | 1 |  |
| Gender |  |  |
| Female | 1.50 | (1.26-1.79) |
| Males | 1 |  |
| Age group (years) |  |  |
| Female |  |  |
| 15-24 | 0.59 | (0.51-0.68) |
| 25-34 | 0.89 | (0.78-1.01) |
| 35-44 | 1.06 | (0.94-1.20) |
| 45-54 | 1.27 | (1.13-1.43) |
| 55-64 | 1.20 | (1.07-1.35) |
| 65-74 | 1.14 | (1.00-1.29) |
| 75- | 1 |  |
| Male |  |  |
| 15-24 | 0.61 | (0.50-0.74) |
| 25-34 | 0.84 | (0.71-1.00) |
| 35-44 | 0.96 | (0.81-1.13) |
| 45-54 | 1.28 | (1.09-1.50) |
| 55-64 | 1.28 | (1.08-1.50) |
| 65-74 | 1.14 | (1.00-1.29) |
| 75- | 1 |  |
| Calendar year |  |  |
| 1997 | 0.89 | (0.80-0.98) |
| 1998-1999 | 0.86 | (0.79-0.93) |
| 2000-2001 | 0.90 | (0.83-0.98) |
| 2002-2003 | 0.88 | (0.81-0.95) |
| 2004-2005 | 0.93 | (0.86-1.00) |
| 2006-2007 | 0.91 | (0.84-0.98) |
| 2008-2009 | 1 |  |
| Urbanicity at birth |  |  |
| Capital | 1.13 | (1.06-1.20) |
| Capital Suburb | 1.05 | (0.95-1.16) |
| Provincial City | 1.25 | (1.16-1.34) |
| Provincial Town | 1.07 | (1.01-1.13) |
| Rural Area | 1 |  |
| Family history of SZ (Maternal) | 1.88 | (1.40-2.54) |
| Family history of SZ (Paternal) | 2.50 | (1.77-3.54) |
| Family history of BP (Maternal) | 6.88 | (5.84-8.10) |
| Family history of BP (Paternal) | 5.64 | (4.55-6.99) |
| Family history of diabetes (Maternal) | 0.99 | (0.88-1.11) |
| Family history of diabetes (Paternal) | 1.09 | (0.98-1.21) |
| History of alcohol abuse | 11.81 | (11.18-12.47) |

<sup>a</sup>Variables are mutually adjusted.
SZ, schizophrenia; BP, bipolar disorder; RR, relative risk

The Diabetes Registry does not subdivide diabetes types 1 and 2, but the Danish National Registry of Patients does make that distinction. Therefore we further analyzed the data as follows: 1) we excluded diabetic patients who had a diagnosis of type 1 in the Danish National Registry of Patients and calculated the risk of type 2 diabetes in patients with SZ and BP: 2) then we examined the risk of developing SZ or BP in patients with each type of diabetes based on the Danish National Registry of Patients. These additional analyses did show the bi-directional association between major mental disorders and type 2 diabetes in the same cohort (**Supplemental Tables 4-6**). Therefore, our main results shown above likely reflect an association between type 2 diabetes and major mental illnesses. From a biological viewpoint, this epidemiological data suggest the existence of common pathophysiological mediators in both diabetes and major mental illnesses.

### Evidence of aberrant insulin signaling and insulin resistance in olfactory neuronal cells biopsied from patients with major mental illnesses (SZ and BP)

As described above, we successfully observed a bi-directional relationship between diagnoses with type 2 diabetes and major mental illnesses in the same cohort (**Fig. 1**). Thus, we hypothesized that common mediators or drivers might underlie both metabolic and mental disorders to explain the bi-directional relationship. To address this question, we used olfactory neuronal cells biopsied from living patients with SZ and BP, as well as those from healthy controls. Olfactory neuronal cells may be a good surrogate tissue to capture neuronal molecular/biological changes associated with disease “pathophysiology”, because these cells are directly used after biopsy from patients and healthy controls without any genetic manipulation or reprogramming [18-20, 30-32]. Olfactory neuronal cells are prepared near to homogeneity [19] and expected to reflect molecular signatures of “pathophysiology”, a converged outcome of genetic and environmental interactions as well as associated stressors [18, 19, 32]. These cells are different from iPS cells from which we expect disease- and individual-associated trait changes (e.g., genetic impacts) related to the disease “pathogenesis” on cellular phenotypes. In the present study, we used two independent sets of data from olfactory neuronal cells.

First, we used the RNA-seq expression profile dataset provided by PsychENCODE [18, 31]. In the study, 36 genes were underscored as differentially expressed (FDR < 0.05) between SZ patients and healthy controls. According to the criteria described in the Method section, we annotated the differentially expressed genes and non-differentially expressed genes into either insulin-related or -unrelated. Among 36 differentially expressed genes, 4 genes (11%) were insulin-related, whereas only 358 genes (1.5%) were insulin-related out of 23,526 genes whose expressions were not significantly different between SZ patients and healthy controls (**Supplemental Table 7**). Thus, in differentially expressed genes between SZ and healthy controls, a significant enrichment of insulin-related genes was indeed validated in olfactory neuronal cells (P = 5.4E-05). These data further support the idea that aberrant insulin signaling may be involved in the pathophysiology of SZ.

Second, we further hypothesized that aberrant insulin signaling may be represented by a hallmark of insulin resistance in olfactory neuronal cells in major mental illnesses, such as SZ. Tyrosine phosphorylation of IRS2 is regarded as a key molecular signature to represent the integrated insulin response [33, 34]: reduction in the level of tyrosine phosphorylation of IRS2 in response to insulin is a well-known indicator of insulin resistance, and a fundamental pathophysiology of type 2 diabetes [35]. Meanwhile, insulin/IRS2 play a key role in cognitive and mental function, in conjunction with their downstream signaling cascades, such as Akt [36, 37]. As far as we are aware, no direct study of IRS2 phosphorylation has been performed using tissue from patients with SZ and BP. Thus, by using olfactory neuronal cells we asked whether a reduction in IRS2 phosphorylation might underlie both diabetes and major mental illnesses (SZ and BP). In this protein study, we used olfactory neuronal cells that were collected at the Johns Hopkins Schizophrenia Center (**Supplemental Table 8;** also see the Method section).

To examine the cellular response of olfactory neuronal cells to insulin, cells were stimulated with insulin and the relative level of the specific phosphorylation of IRS2 to the overall expression of IRS2 protein was assessed by immunoprecipitation followed by Western blotting (IP-Western) (**Supplemental Fig. 1**) As shown in **Fig. 2**, insulin-stimulated tyrosine phosphorylation of IRS2 was significantly decreased in SZ patient-derived olfactory neuronal cells (about 50% of controls). A consistent trend of reduction was also found in BP patient-derived cells (about 75% of controls). Furthermore, we excluded one SZ patient who was pre-diabetic and 2 BP patients who had diabetes at the time of the nasal biopsy, and conducted the same analysis: the new results without these 3 pre-diabetic and diabetic subjects lead to the same conclusions as those we obtained from the analysis that included all subjects (**Supplemental Fig. 2**). Removal of one possible outlier (higher than the average at more than two-fold standard deviation) from the healthy control group did not affect the significant difference between SZ patients and healthy controls (**Supplemental Fig. 3**). The fact that neuronal cells derived from patients with SZ or BP responded poorly to insulin stimulation, a characteristic also observed in subjects with insulin resistance, suggested that common molecular mediators involving insulin/IRS2 signaling might underlie both diabetes and major mental illnesses.

**Fig. 2.**
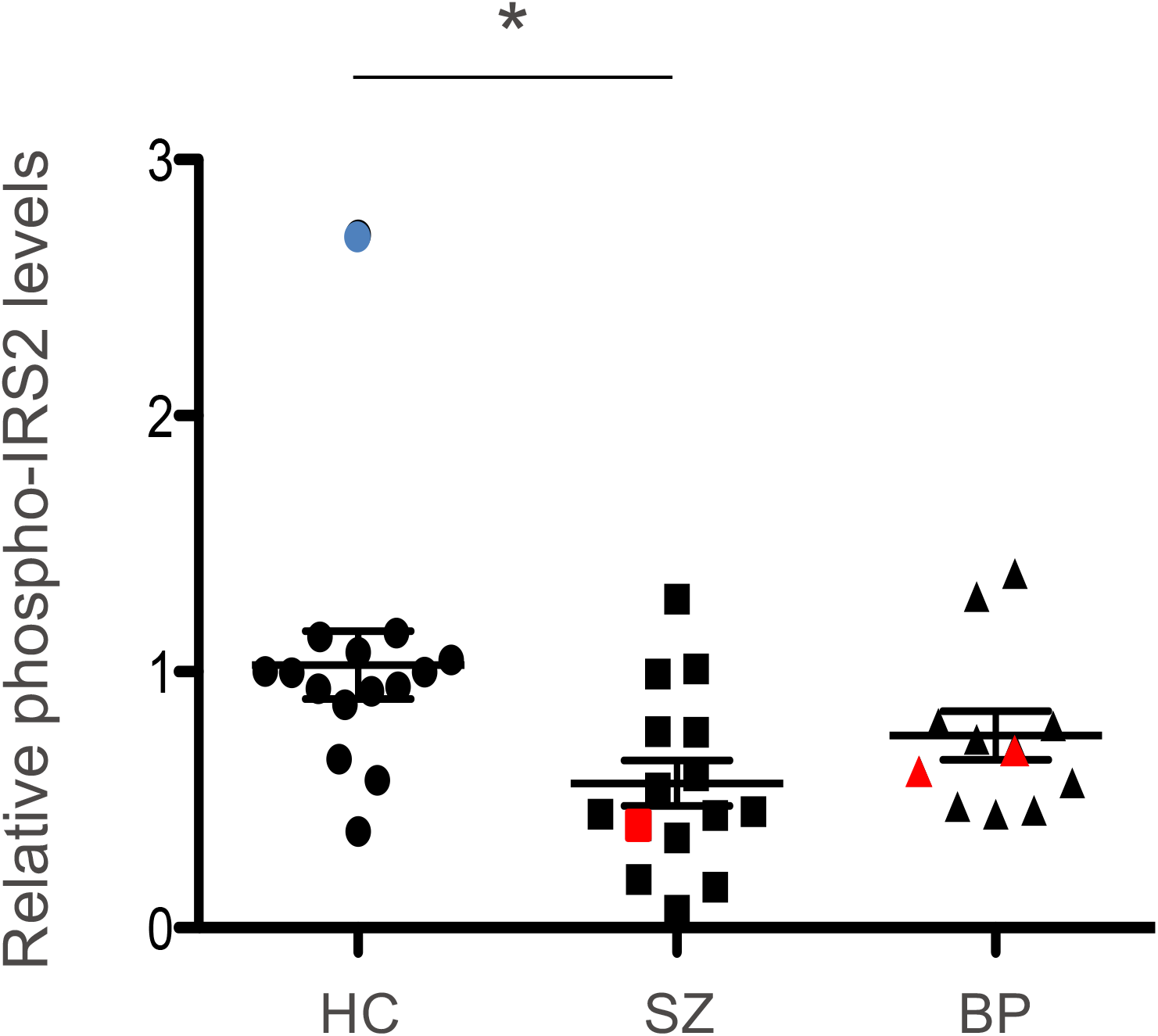
Tyrosine phosphorylation of IRS2 in olfactory neuronal cells derived from SZ and BP patients. Olfactory neuronal cells derived from patients with SZ or BP as well as those from healthy controls (HC) were stimulated with insulin and characterized for the level of tyrosine phosphorylation of IRS2. Data are shown as mean ± SEM. * P<0.013. Red, 1 SZ patient who was pre-diabetic and 2 BP patients who had diabetes at the time of the nasal biopsy. Blue, an outlier that is different from the average at more than two-fold the standard deviation. Notes: the data in the case when we removed the 3 patients in pre-diabetic or diabetic condition are shown in **Supplemental Fig. 2**. Furthermore, the data in the case we removed the outlier are shown in **Supplemental Fig. 3**. In both cases, the scientific conclusions are the same.

A previous report with animal models indicated that phosphorylation of IRS1 at serine 307 was upregulated in an insulin resistance condition at baseline (without insulin stimulation) *in vivo* [38]. Thus, we tested the phosphorylation of serine 312 (the equivalent residue in humans to mouse serine 307) and observed that IRS1 phosphorylation at serine 312 was upregulated at baseline (without insulin stimulation) in olfactory neuronal cells from SZ patients, compared with controls (**Supplemental Fig. 4**), whereas no difference was observed between BP patients and controls. Taken together, molecular signatures of insulin resistance were observed in both IRS1 and IRS2 in neuronal cells from SZ patients, and those also observed at least in IRS2 in neuronal cells from BP patients.

### Dopamine-associated behavioral changes in *Irs2* knockout (KO) mice

Next we explored these putative common mediators (e.g., IRS2 and IRS1) in higher brain function relevant to major mental illnesses by using a genetically engineered model. As the first step of a long-term effort in addressing the mechanism, we decided to address a loss of function of IRS2. We selected IRS2 as the first target since IRS2 is more thoroughly studied in the brain compared with IRS1 [20, 36, 37, 39]: nevertheless, its investigation has not yet extended to the biology of mental disorders. Thus, we examined behavioral changes relevant to major mental illnesses in *Irs2* knockout mice (KO). In order to avoid the potential complications of premature death due to excessive hyperglycemia [35, 40], we used male KO animals at 7 weeks of age (late adolescence to young adulthood) for our analysis. Due to deficits in motor function observed in the rotarod paradigm (**Supplemental Fig. 5a**), the interpretation of experimental results was not straightforward. For example, KO mice showed decreased spontaneous activity in the open field, as well as total distance in the elevated plus maze and Y-maze tasks, which may possibly be associated with motor deficits (**Supplemental Fig. 5b-d**). We did not observe any significant difference between WT and KO mice in the three chamber social interaction tasks (**Supplemental Fig. 5e**). In contrast, *Irs2* KO mice showed increased immobility in the forced swim test (**Supplemental Fig. 5f**). This finding may be indicative of depression-associated changes, consistent with human findings that individuals with diabetes have an elevated risk for depression [41]. Nevertheless, we cannot fully exclude the possibility that this result is due to impaired motor function.

Because the deficits in motor function are a potential confound in the interpretation of the data, we introduced a challenge study using the psychostimulant, methamphetamine, to address neurochemical changes *in vivo*. Patients with psychotic disorders are known to show hypersensitivity to psychostimulants such as methamphetamine [42], and such pathophysiology has been studied in mouse models as a relative increase in locomotion compared to baseline (before psychostimulant administration) [43]. Interestingly, KO mice showed a significantly greater response to methamphetamine compared to WT mice (**Fig. 3**). We observed two peaks of augmented responses to methamphetamine in KO mice, which is not usually seen in WT mice.

**Fig. 3.**
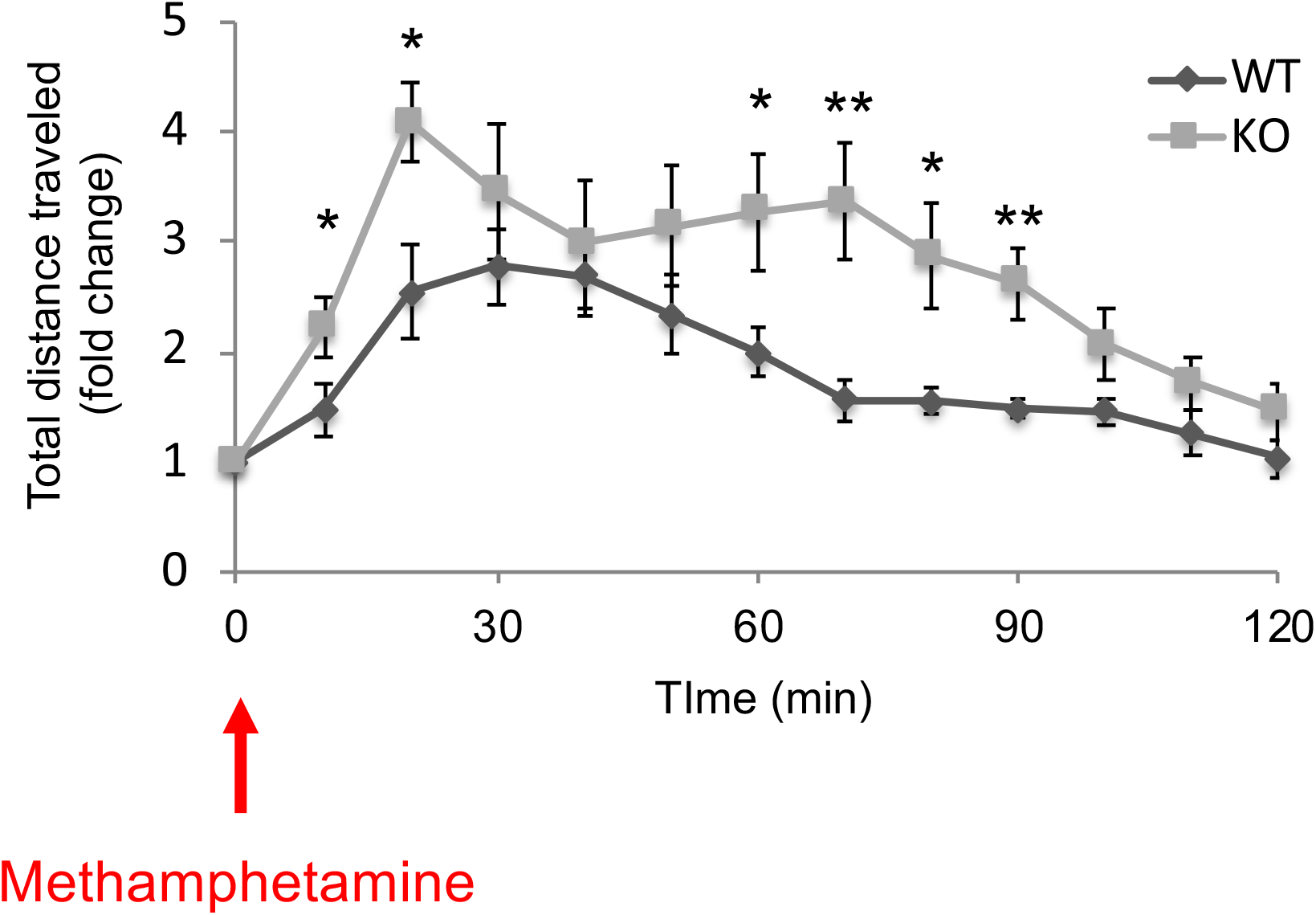
Response of *Irs2* knockout mice after methamphetamine administration. A methamphetamine challenge test was conducted in 10 wild type (WT) and 9 knockout (KO) mice. The fold change in the total distance traveled every 10 min after methamphetamine injection relative to baseline activity is demonstrated. Data are shown as mean ± SEM. * P<0.05, ** P<0.01.

## DISCUSSION

In the present study, we demonstrated a higher risk of developing major mental illnesses in patients who had already been diagnosed with diabetes compared to that in the general population. We also confirmed a well-documented observation that patients who had already been diagnosed with major mental illnesses (SZ or BP) had a higher risk of developing diabetes compared to the general population. As far as we are aware, this is the first time that this bi-directional relationship between the risks for major mental illnesses and diabetes has been demonstrated within the same cohort. This bi-directional relationship observed in high-quality population data from Denmark suggested that there are common pathophysiological mediators or drivers that underlie both mental and physical conditions. Importantly, by demonstrating the cases in which diabetes precedes the onset of major mental illnesses, the results are unlikely to be confounded by the effects of antipsychotic medications. SZ and BP are categorized as two distinct illnesses in the DSM-V (Diagnostic and Statistical Manual of Mental Disorders, Fifth Edition). However, recent studies of human genetics, brain imaging, and neuropsychology have shown that these two diseases share risk factors and phenotypic dimensions [5, 44, 45]. In the present study both SZ and BP show comorbidity with diabetes, which is consistent with the recent reports in genetic, imaging, and cognitive studies.

To explore possible common mediators stemming from the epidemiological data, we took a hypothesis-driven, candidate molecular approach by using both patient-derived neuronal cells and animal models. Olfactory neuronal cells obtained through nasal biopsy from living patients and healthy subjects are expected to reflect molecular signatures of “pathophysiology”, a converged outcome of genetic and environmental interactions as well as associated stressors [18, 19, 32]. Accordingly, we suggest that alteration of insulin/IRS2 signaling underlies, at least in part, both major mental illnesses and diabetes, likely mediating their shared pathophysiology. If metabolic problems such as insulin resistance and mental illnesses share common pathophysiological mediators, treatments for metabolic problems, including diabetes or insulin resistance, may serve as a supplement or alternative to antipsychotics. For instance, the indirect antioxidant sulforaphane (SFN), a broccoli-derived phytochemical now commercially available in concentrated dietary supplements, reportedly ameliorates the pathophysiology of obese patients with dysregulated type 2 diabetes [46]. In parallel, in psychiatry research, the application of SFN to mental disorders is considered with promising results [47-52]. We may also be able to use glucose intolerance as a biomarker for stratification of patient populations for possible treatment by metabolic correction, as well as for monitoring the effectiveness of the treatment.

Our aspiration in the present study is to connect an epidemiological study with molecular investigation, with the latter inspired by the former. We acknowledge that this ambitious approach may inevitably receive questions or criticisms when we reduce epidemiological information (macroscopic) to molecular targets/pathways (microscopic). To minimize questions and criticisms on reductionism and disconnection, it is important to choose molecular targets in disease pathophysiology or acquired phenotypes (not each single etiology or pathogenesis) that are expected to represent a converged outcome of genetic and environmental interactions as well as associated stressors [53]. We believe that phosphorylation of IRS proteins for insulin resistance is a good example (even if not perfectly ideal) that satisfies this expectation [54-58], like phosphorylation of tau in multiple neurodegenerative disorders [59, 60]. Based on the evidence of IRS phosphorylation as a promising and solid entry point, we will be able to develop fruitful biology that bridges both metabolic and mental conditions. For example, although it is beyond the scope of the present study, investigating the whole genome epigenetic landscape in olfactory neuronal cells from SZ and BP patients may deepen the biology by using IRS phosphorylation changes as a landmark.

This study includes other points that are to be carefully considered. First, as noted above, the diabetic population in this study had a relatively short duration of diabetes, and may not necessarily represent the general population of patients with diabetes. Second, since the Diabetes Registry does not subdivide diabetes types 1 and 2, we could not completely exclude the possibility that the subtypes were a confounding factor: however, the additional analysis which used the subdivision of diabetes based on the Registry of Patients suggest that our results reflect the association between type 2 diabetes and major mental illnesses. Third, we were not able to adjust for some factors that may have confounded the results, such as the usage of antipsychotics or obesity (e.g., body mass index). Indeed, antipsychotic-related risks for diabetes in young populations have been shown in the literature [61]. On the other hand, glucose intolerance or insulin resistance has been observed even in antipsychotic-naïve patients [12-14]. Together, we believe that our finding (i.e., high risk of developing diabetes in SZ/BP patients) is not merely a consequence of antipsychotic medications. Furthermore, as described above, the finding that diabetes precedes the onset of major mental illnesses (shown in **Fig. 1b**) suggests an intrinsic link of diabetes and major mental illnesses beyond the medication effects. Fourth, we were not able to include factors associated with lifestyle such as smoking, diet, and exercise habits due to the absence of standardized variables representing life style factors in the Danish cohorts, or factors such as socio-economic status and income level that might be associated with risk for mental disorders and diabetes. Fifth, although our hypothesis-driven candidate molecule approach focusing on IRS2 provided evidence supporting IRS2 as a common mediator that underlies both major mental illnesses and diabetes, we did not exclude the possibility that there are other common mediators for these conditions and such other mediators will be explored in future studies in an unbiased approach. Sixth, although in the present study we simply tested behavioral impact elicited by a loss of function of IRS2, future studies will include different mouse models with a wider range of behavioral assays.

The present study provides a unique and novel platform to study the mechanisms of psychiatric conditions by combining 1) classical epidemiological study, 2) study with biospecimens from patients and controls, and 3) study of animal models. High-quality population-based epidemiological analysis can aid in developing hypotheses that can be tested in human tissues and animal models. In human tissues, we can test the clinical validity of epidemiology-driven working hypotheses at the molecular level. However, to address the causality of the molecular changes to behaviors, we would need to include animal models in combination with human tissues in the study design. In the present study, by using these experimental resources, we proposed a common molecular signature, i.e., reduction in the tyrosine phosphorylation of IRS2, between major mental illnesses (SZ and BP) and diabetes. In future studies, instead of the candidate molecular approaches adopted here, we may compare molecular signatures in tissues between patients with major mental illnesses and those with diabetes in an unbiased manner, and further validate promising common signatures using relevant animal models. The approach of “from population to neuron” used in the present study may be a promising strategy to decipher clinically relevant molecular mechanisms and a pathway leading to possible new therapeutic approaches.

## Data Availability

Lead contact: Akira Sawa.

## ACKNOWLEDGEMENTS

We thank Yukiko Y. Lema for organizing the manuscript and figures, Drs. Melissa A. Landek-Salgado and Nao J. Gamo for critical reading of the manuscript. AS and KI are supported by the National Institute of Mental Health MH-105660 and Maryland Stem Cell Research Fund. AS is also supported by MH-094268 Silvio O. Conte center, MH-092443, and MH-107730, as well as foundation grants from Stanley, S-R/RUSK, BBRF. TS is supported by Kakenhi from the Ministry of Education, Culture, Sports, Science and Technology in Japan, and was funded in part by the Takeda and Kyoto University Basic and Clinical Research Project for CNS Drugs (supported in part by Takeda Pharmaceutical Co. Ltd.). TML was supported by an unrestricted grant from the Stanley Medical Research Institute. PBM was supported by grants from the Danish Strategic Research Council, the Faculty of Social Sciences at Aarhus and a European Research University, the Lundbeck Foundation, the Stanley Medical Research Institute, Council advanced grant (GA 2948338). WWE was supported by NIMH grant 1R34MH007760-01.

## SUPPLEMENTAL FIGURE LEGENDS

**Supplemental Fig. 1 | Tyrosine phosphorylation of IRS2 in olfactory neuronal cells after insulin stimulation** Olfactory neuronal cells derived from healthy controls were stimulated with different concentrations of insulin and characterized for the level of tyrosine phosphorylation of IRS2 (pIRS2). In this experiment, we demonstrated that the linearity of the signal (the level of tyrosine phosphorylation of IRS2) occurs between the insulin concentrations of 0 and 30 nM, which is indicated by a dotted line on the graph. Thus, we used 10 nM insulin for all experiments to be within the linear range of the pIRS2 signal.

**Supplemental Fig. 2 | Tyrosine phosphorylation of IRS2 in olfactory neuronal cells after excluding pre-diabetic and diabetic patients from the analysis (see Figure 2 including all the patients)** Olfactory neuronal cells derived from patients with SZ or BP as well as those from healthy controls (HC) were stimulated with insulin and characterized for the level of tyrosine phosphorylation of IRS2. Data are shown as mean ± SEM. * P=0.0133 (one-way ANOVA, followed by Bonferroni’s multiple comparison test). Though one SZ patient who was pre-diabetic and 2 BP patients who had diabetes at the time of the nasal biopsy were excluded from this analysis, the results did not change from the analysis including all the patients.

**Supplemental Fig. 3 | Tyrosine phosphorylation of IRS2 in olfactory neuronal cells after excluding one healthy control outlier from the analysis (see Figure 2 including all the subjects)** Olfactory neuronal cells derived from patients with SZ or BP as well as those from healthy controls (HC) were stimulated with insulin and characterized for the level of tyrosine phosphorylation of IRS2. Data are shown as mean ± SEM. * P=0.0269 (one-way ANOVA, followed by Bonferroni’s multiple comparison test). Though one possible outlier (higher than the average by more than two-fold standard deviation) was excluded in this analysis, the results did not change from the analysis including all the patients.

**Supplemental Fig. 4 | IRS1 phosphorylation at serine-312 in olfactory neuronal cells derived from SZ and BP patients** The level of IRS1 phosphorylation at serine-312 (pIRS1) was measured in olfactory neuronal cells derived from patients with SZ or BP and healthy controls (HC) at baseline (without insulin stimulation). Data are shown as mean ± SEM. *P<0.05. Red, 1 SZ patient who was pre-diabetic and 2 BP patients who had diabetes at the time of the nasal biopsy. Green, 2 outliers (1 SZ and 1 BP) that are different from the average at more than two-fold standard deviation. The inclusion and exclusion of these subjects (diabetic/pre-diabetic patients or outliers) did not change the scientific conclusions.

**Supplemental Fig. 5 | Behavioral assays of *Irs2* KO mice** Male *Irs2* KO mice (N=9) and WT littermates (N=12) at 7 weeks of age were subjected to rotarod (a), open field (b), elevated plus maze (c), Y-maze (d), three-chamber social interaction (e), and forced swim test (f). Data are shown as mean ± SEM. Statistical analysis was performed as described in the Materials and Methods. * P<0.05, ** P<0.01, *** P<0.001.

